# Risk compensation after HIV-1 vaccination may accelerate viral adaptation and reduce cost-effectiveness: a modeling study

**DOI:** 10.1101/2020.12.13.20248146

**Authors:** Kathryn Peebles, John E. Mittler, Steven M. Goodreau, James T. Murphy, Molly C. Reid, Neil Abernethy, Geoffrey S. Gottlieb, Ruanne V. Barnabas, Joshua T. Herbeck

## Abstract

Pathogen populations can evolve in response to selective pressure from vaccine-induced immune responses. For HIV, models predict that viral adaptation, either via strain replacement or selection on *de novo* mutation, may rapidly reduce the effectiveness of an HIV vaccine. We hypothesized that behavioral risk compensation after vaccination may accelerate the transmission of vaccine resistant strains, increasing the rate of viral adaptation and leading to a more rapid decline in vaccine effectiveness. To test our hypothesis, we modeled: a) the impact of risk compensation on rates of HIV adaptation via strain replacement in response to a partially effective vaccine; and b) the combined impact of risk compensation and viral adaptation on vaccine-mediated epidemic control. We used an agent-based epidemic model that was calibrated to HIV-1 trends in South Africa, and includes demographics, sexual network structure and behavior, and within-host disease dynamics. Our model predicts that risk compensation can increase the rate of HIV viral adaptation in response to a vaccine. In combination, risk compensation and viral adaptation can, under certain scenarios, reverse initial declines in prevalence due to vaccination, and result in HIV prevalence at 15 years equal to or greater than prevalence without a vaccine.

## Introduction

Recent years have seen increased availability of effective biomedical HIV-1 prevention tools, including the provision of antiretroviral therapy (ART) for prevention and pre-exposure prophylaxis (PrEP). However, challenges in ART coverage and uptake of PrEP indicate that an HIV-1 vaccine will be a necessary addition to combination prevention for epidemic control.^1^ To date, one HIV-1 vaccine trial (RV144) has shown modest efficacy, reducing the risk of HIV-1 infection by 31% at 3.5 years after vaccination.^2^ However, post-hoc sieve analyses^3^ showed that the vaccine was differentially efficacious against viral genotypes, with efficacy ranging from 40% to 80% against three viral genotypes and a non-significant effect against others.^4^ These results suggest that the observed 31% vaccine efficacy (reduction in overall rate of infection) was an aggregate over viral genotypes present in the population; i.e., this population level vaccine efficacy estimate is an average of strain-specific efficacies, weighted by strain population frequency. Our previous work predicted that HIV-1 viral diversity may lead to viral adaptation in response to a partially effective leaky vaccine, reducing vaccine effectiveness at the population level over time as resistant strains rise in frequency.^5^ This process of vaccine-induced viral adaptation (via strain replacement) has been empirically observed in other pathogens, including *Streptococcus pneumoniae, Haemophilus influenzae*, and others.^6,7^

Risk compensation may pose an additional threat to population-level impact of a partially effective vaccine.^8^ Risk homeostasis theory posits that each individual has a unique risk set point, with the consequence that reductions in risk achieved by one preventive therapy or behavior will be offset by increases in risk through compensatory changes in behavior to maintain an equilibrium risk level.^9^ In a study of HIV vaccine attitudes among high-risk adults in Los Angeles, 10% of respondents indicated they would reduce condom use if they were vaccinated, and this risk compensation was positively associated with perceived acceptability of a potential vaccine.^10,11^ In another study of vaccine acceptability and risk compensation among high-risk MSM and transgender people in Thailand, over one-third of participants reported intentions to increase post-vaccination risk behaviors.^12^ In a network-based health decision model of vaccination for HIV in a theoretical population, unprotected sex increased after vaccination.^13^ Observational studies of risk compensation for human vaccines that have already been introduced are mixed: no increases in high risk behavior were identified among HPV vaccinated individuals (reviewed in Brewer^14^).

Observational data from other HIV prevention measures suggest that risk compensation may occur with a preventative HIV vaccine: as PrEP has become widely available, evidence from demonstration projects and implementation settings in high-income countries has shown an increase in condom-less anal sex among PrEP users,^15-18^ although no increase in the number of sexual partners has been observed.^17,18^ An increase in incidence of sexually transmitted infections among PrEP users relative to non-users also provides indirect evidence of risk compensation.^19,20^ This might not apply generally, as no evidence of risk compensation following PrEP initiation was seen for heterosexual HIV serodiscordant couples in Kenya and Uganda.^21^ Evidence from sub-Saharan African settings of risk compensation in response to rollout of voluntary medical male circumcision is also mixed, with two of three of the randomized clinical trials finding evidence of small increases in condom-related and sexual contact frequency risk behaviors,^22-24^ but not with respect to number of partners, and recent evidence following rollout has not identified significant increases in risk behaviors among circumcised men.^25-29^ An observational study in Kenya estimated that men undergoing voluntary medical circumcision were 42% more likely to report two or more sexual partners in the previous year.^30^

Previous mathematical modeling of partially effective HIV-1 vaccines similar to that tested in RV144 has predicted that a vaccine with 60% coverage will avert 6-15% of infections over 10 years,^31-33^ with greater gains achieved with continuous coverage and regular booster vaccination. A review of models using similar methods to assess the cost-effectiveness of a partially effective HIV-1 vaccine concluded that an HIV-1 vaccine would be a cost-effective prevention strategy under the majority of conditions and assumptions, with potential cost savings with targeted vaccination strategies.^33,34^

However, these analyses have largely overlooked the mitigating factors of viral adaptation and risk compensation, and none have modeled both factors simultaneously. Previous models have predicted a limited effect of risk compensation on vaccine impact, with large reductions in incidence even in the presence of increases in risky behavior among vaccinated individuals.^31,32,35,36^ Yet increased risk behavior by vaccinated individuals, under the assumption of protection, may result in more frequent transmission of resistant viruses and lead to the faster spread of vaccine resistance and a more rapid decline in vaccine effectiveness.

Here we evaluate the potential combined effect of risk compensation and viral adaptation through epidemic and evolutionary modeling of an HIV-1 vaccine implemented in South Africa. We hypothesize that risk compensation will accelerate rates of viral adaptation, resulting in a larger detrimental impact on vaccine effectiveness than viral adaptation and risk compensation operating alone. We further hypothesize that viral adaptation and risk compensation will reduce the cost-effectiveness of a vaccine.

## Results

Using a stochastic, agent-based HIV-1 epidemic model calibrated to South Africa, we modeled: 1) the impact of behavioral risk compensation on rates of viral population-level adaptation in response to vaccination; and 2) the combined effects of risk compensation and viral adaptation on vaccine-mediated epidemic control. We also calculated the cost-effectiveness of a partially effective HIV vaccine accounting for viral adaptation and risk compensation.

In the absence of both vaccine-resistant viral strains and risk compensation, our baseline model scenario predicts that an HIV-1 vaccine with 50% efficacy implemented with 70% coverage may result in an HIV-1 prevalence of 7.8% (95% CrI: 7.0, 8.4), following 15 years of continuous vaccination, compared to prevalence of 13.6% (95% CrI: 12.5, 14.9) in a counterfactual scenario without a vaccine (Figure 1; Table S1). This decline in prevalence reflects 56.8% (95% CrI: 41.4, 69.0) of infections averted over that time span (compared to the counterfactual with no vaccine) (Figure 2; Table S2).

**Figure 1.**
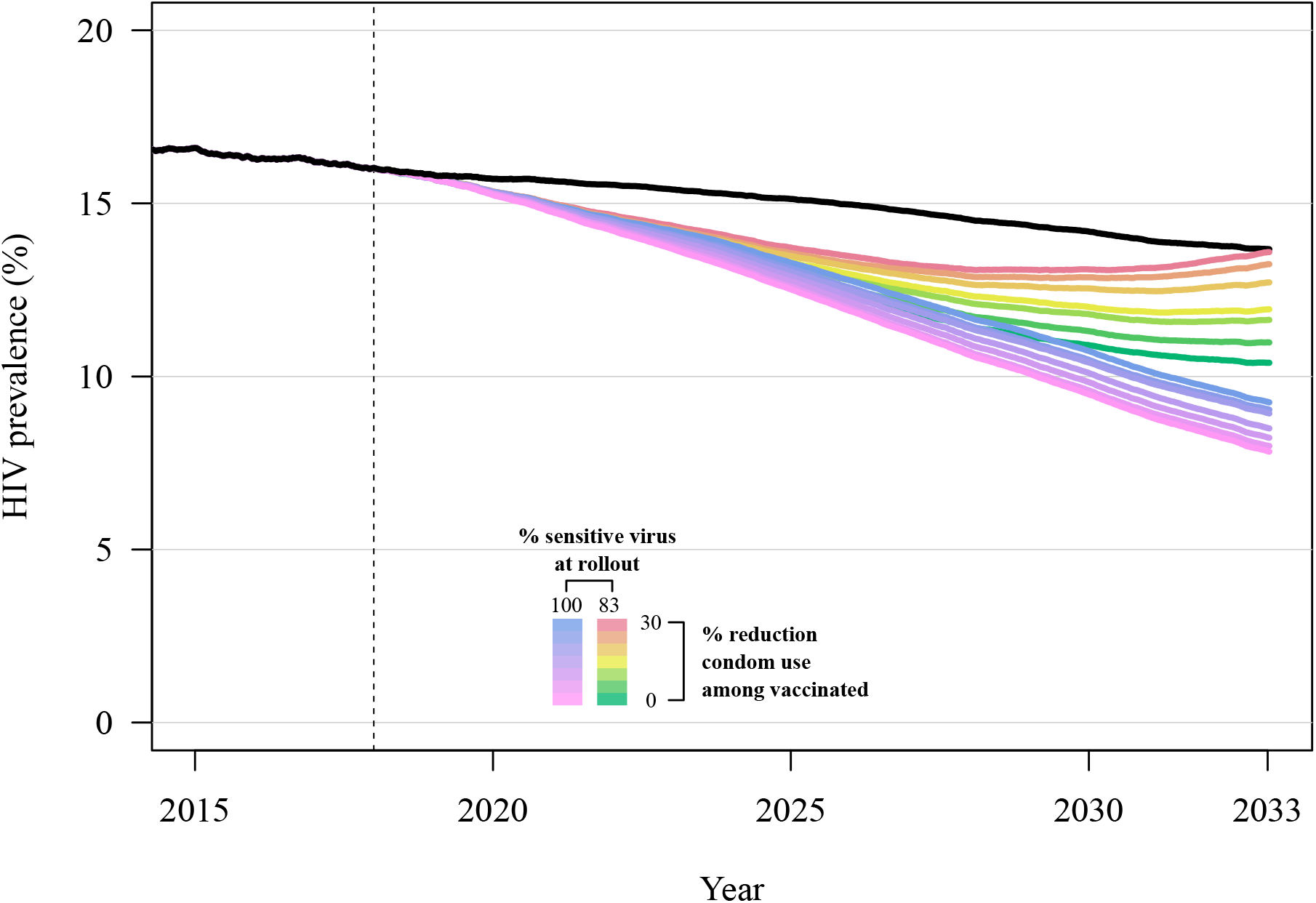
HIV-1 prevalence following introduction of a partially effective vaccine in the year 2018 (dashed vertical line), with 50% overall vaccine efficacy and 70% vaccine coverage. Model scenarios shown include variation in the proportion of vaccine-sensitive virus at vaccine rollout (83% or 100%) and variation in the reduction in condom use among vaccinated individuals (from 0% to 30% reduction). The mean and 95% credible interval of 64 replicates of each scenario are shown. The black line shows the model scenario without a vaccine.

**Figure 2.**
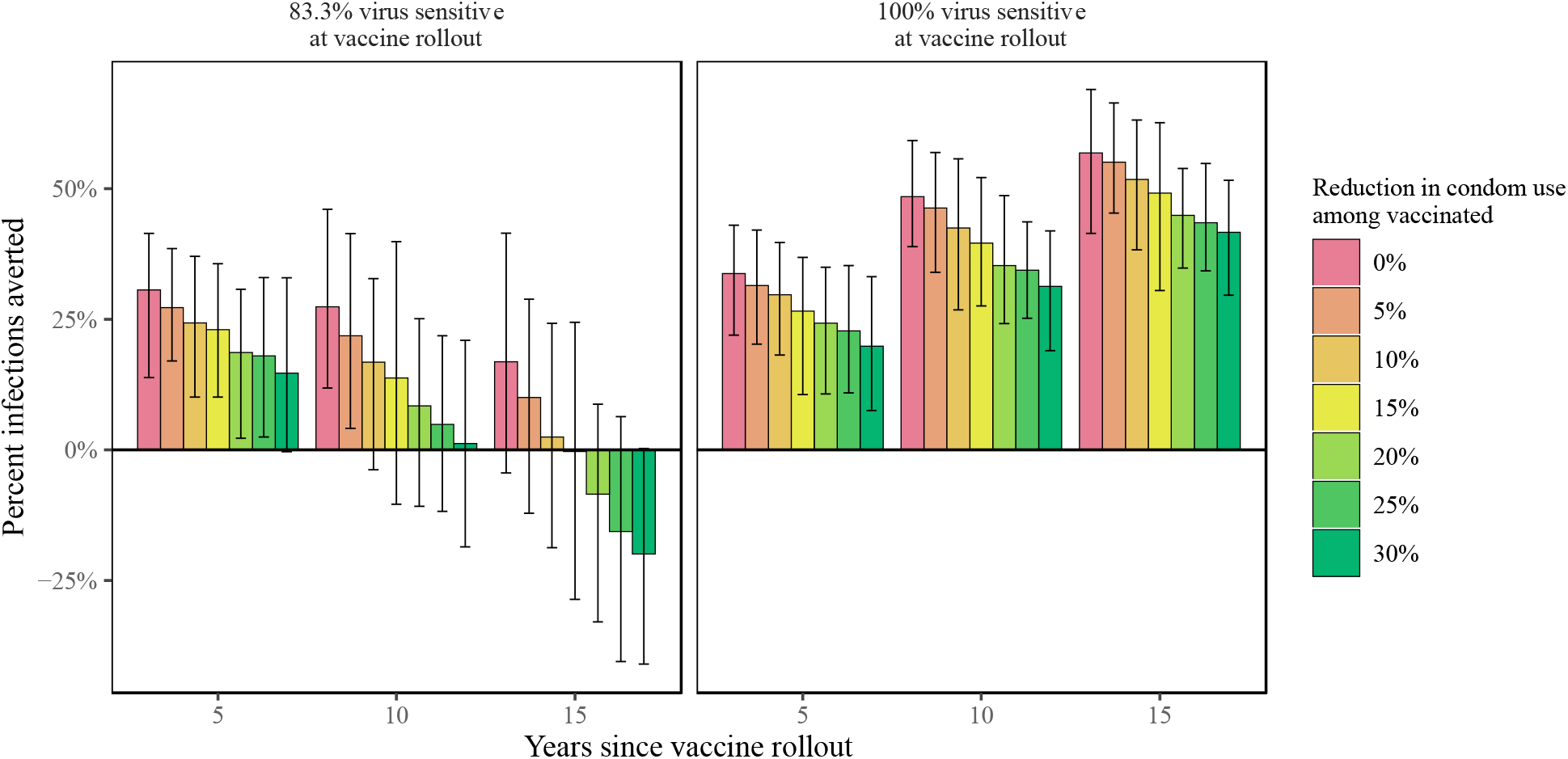
Percent of HIV-1 infections averted via vaccination over the first 15 years after vaccine rollout, in model scenarios that include variation in the proportion of vaccine-sensitive virus at vaccine rollout (83% or 100%) and variation in the reduction in condom use among vaccinated individuals (from 0% to 30% reduction), with 50% overall vaccine efficacy and 70% vaccine coverage.

### Impact of risk compensation on rates of HIV adaptation

For the baseline scenario *with* viral adaptation (50% overall vaccine efficacy, 70% vaccine coverage, and 17% of virus vaccine resistant at rollout) but in the absence of risk compensation, our model predicts that the vaccine resistant allele frequency will increase over time after the rollout of a partially effective vaccine: within 15 years the resistant allele frequency has increased from 17% to ∼45% (Figure 3). Likewise, during this time period the resistant allele incidence has increased from ∼0.4% to ∼1.3% (Figure 4). In scenarios with higher initial frequencies of resistant virus (i.e. 28.6%, 37.5%), the resistant virus frequency reaches ∼60% and ∼70% after 15 years, respectively (Figure S3). With viral adaptation *and* risk compensation, HIV adaptation occurs more rapidly. In the baseline model scenario with adaptation and risk compensation, within 15 years the resistant allele frequency has increased from 17% to ∼55% and incidence has increased from ∼0.4% to ∼2.4% (Figure S3).

**Figure 3.**
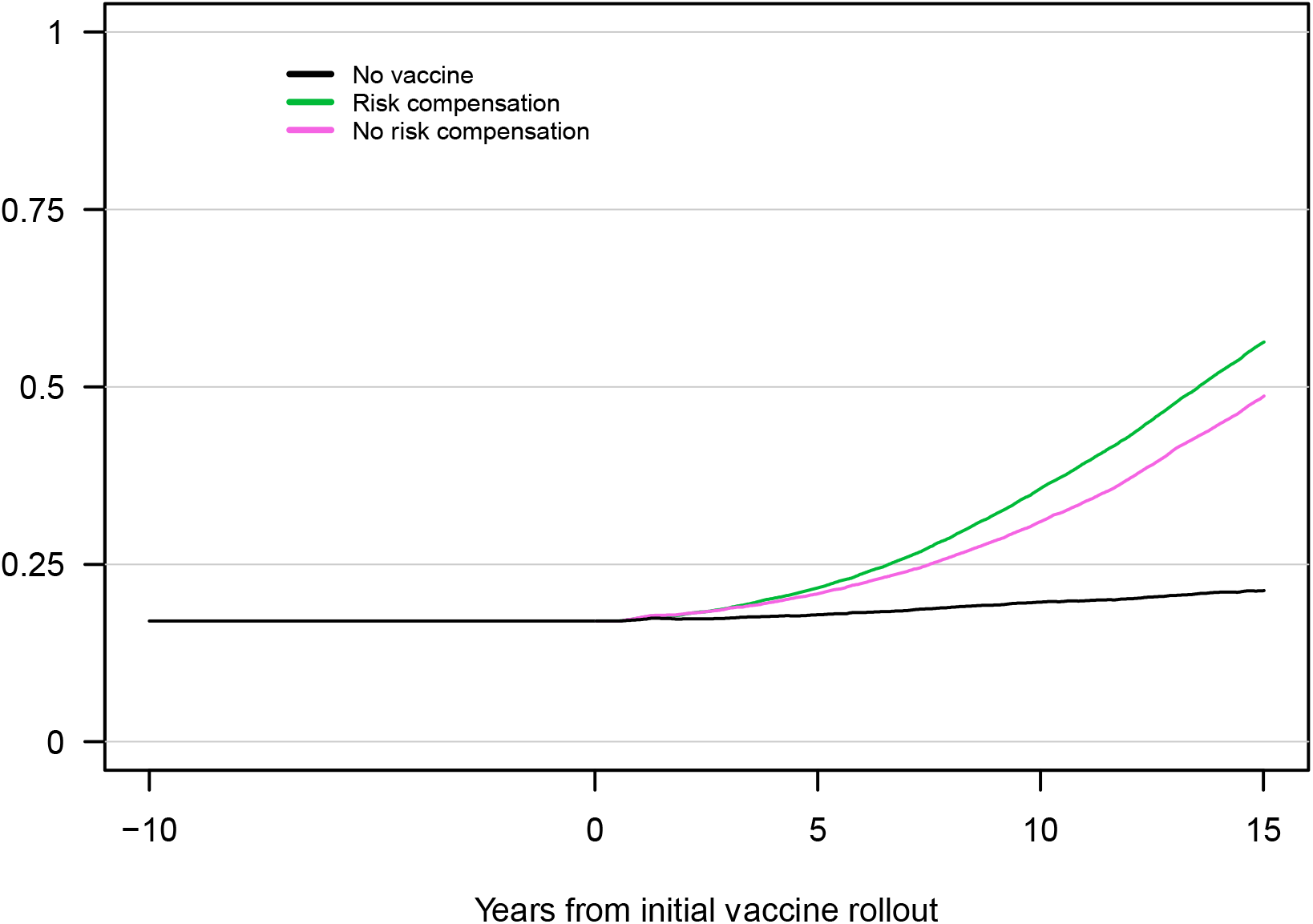
Frequencies of HIV genotypes that are either sensitive or resistant to vaccine induced immune responses, following introduction of a partially effective vaccine (here shown as years from vaccine rollout). The plot shows the mean and 95% credible interval of 64 replicates for each scenario (no vaccine, vaccine with risk compensation, and vaccine without risk compensation; vaccine scenarios 83.3% vaccine sensitive virus at vaccine rollout and 70% population vaccine coverage). Risk compensation is modeled as a 30% reduction in condom use among vaccinated individuals.

**Figure 4.**
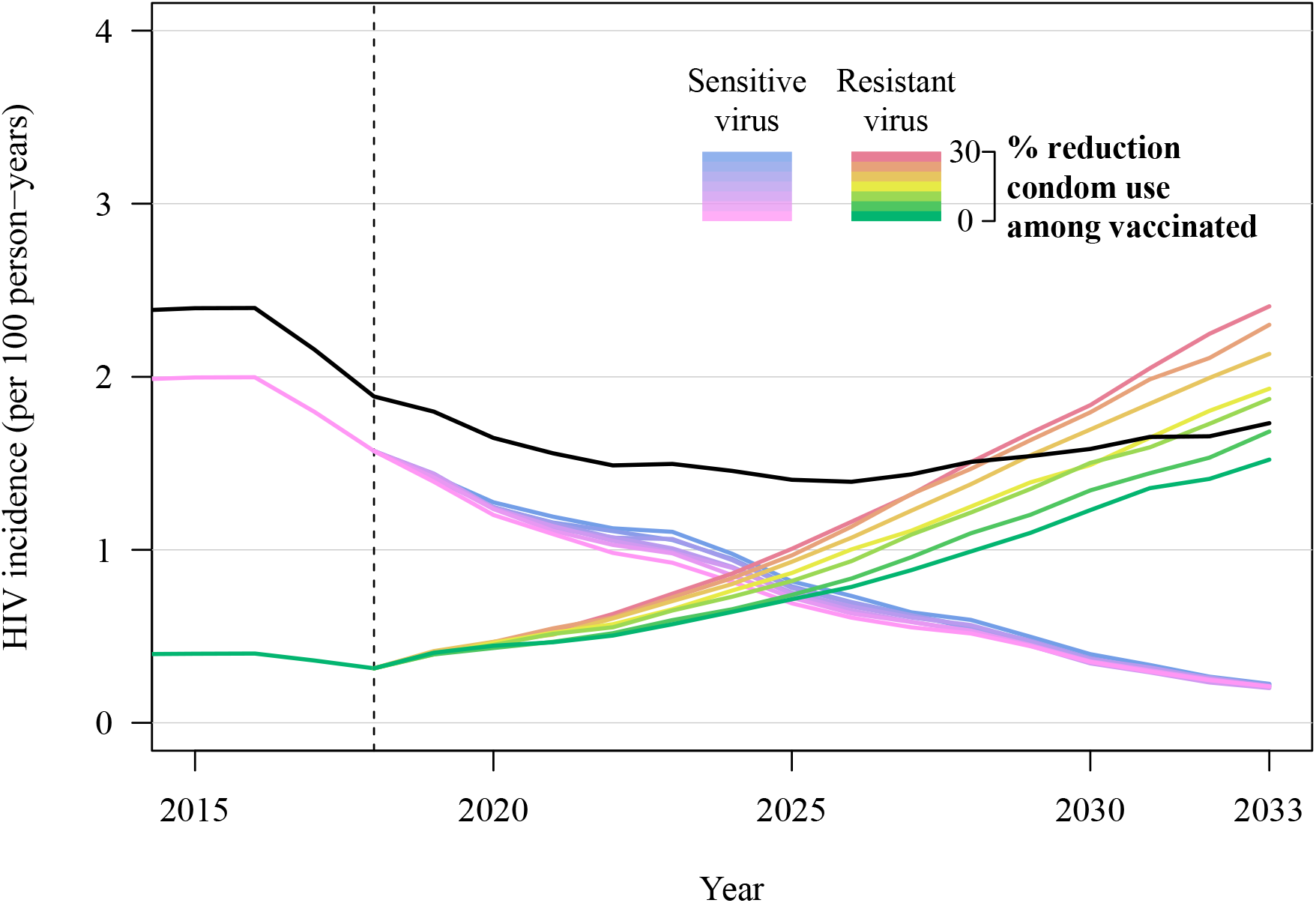
HIV-1 incidence of vaccine-sensitive and vaccine-resistant viral strains following introduction of a partially effective vaccine in the year 2018 (dashed vertical line), with 50% overall vaccine efficacy and 70% vaccine coverage. Model scenarios include variation in the reduction in condom use among vaccinated individuals (from 0% to 30% reduction). The mean and 95% credible interval of 64 replicates of each scenario are shown. Black line shows total incidence (incidence from sensitive and resistant viruses combined).

### Combined impact of risk compensation and HIV adaptation on vaccine effectiveness

With the baseline scenario and risk compensation alone (without vaccine-resistant viral strains), the vaccine is predicted to result in decreased incidence and prevalence, and lead to infections averted, yet with a weaker impact compared to the baseline scenario *without* risk compensation (Figure 1, Figures S4, S5, S6). A 5% decrease in condom use among vaccinated individuals results in 8.0% (95% CrI: 7.3, 8.7) prevalence at fifteen years after vaccine rollout, while a 30% decrease in condom use among vaccinated individuals results in 9.3% prevalence (95% CrI: 8.6, 10.2), both compared to a prevalence of 7.8 (95% CrI: 7.0, 8.4) without risk compensation (reduced from 13.6% prevalence without vaccination). These scenarios result in 55.1% (95% CrI: 42.3, 69.6) and 41.6% (95% CrI: 29.3, 50.5) of infections averted, for 5% and 30% decreases in condom use, respectively, compared to 56.8% of infections averted with no risk compensation (Table S2).

With only the presence of vaccine-resistant viral strains (without risk compensation), with 17% of virus vaccine resistant at rollout, viral population-level adaptation substantially reduces vaccine impact: HIV prevalence is reduced to 10.4% (95% CrI: 8.3, 11.8) within 15 years after vaccine rollout, compared to a reduction to 7.8% prevalence in the baseline scenario in which all virus is vaccine sensitive (Figure 1, Table S1). Initial declines in prevalence are driven by rapid decreases in incidence of vaccine-sensitive viral strains; the near elimination of vaccine-sensitive viral infections is accompanied by an increase in the incidence of vaccine-resistant infections, which represent the majority of incident viral strains within seven years of vaccine rollout (Figure 4). In the same comparison of scenarios, infections averted through vaccination (15 years after rollout) declined from 56.8% averted in the baseline scenario *without* resistant virus to only 16.8% averted (95% CrI: −4.4, 41.5) with 17% resistant virus at rollout (Figure 2). Most notable, HIV-1 prevalence following fifteen years of vaccination increases (prevalence range: 10.4%-11.8%) as vaccine coverage and initial resistant virus frequency increase (Figure 1, Table S1).

The presence of *both* risk compensation and viral adaptation results in poorer epidemic control than with either operating alone. As described above, compared to the baseline scenario, risk compensation alone (via a 30% reduction in condom use among vaccinated individuals and no viral adaptation) may reduce the vaccine impact on prevalence by 1.5 percentage points and avert 17.2% fewer infections; viral adaptation alone (with no risk compensation) may reduce the vaccine impact on prevalence by 2.6 percentage points and avert 40% fewer infections. Yet with both risk compensation and viral adaptation, HIV prevalence is 5.8 percentage points higher than in a scenario with neither risk compensation nor viral adaptation (Figure 1, Table S1), an absolute difference of 1.8 percentage points above the simple additive effect of viral adaptation and risk compensation. In this scenario, initial declines in prevalence are reversed at approximately 10 years following vaccine rollout, as the combination of risk compensation and viral adaptation result in a rebound in prevalence such that following 15 years of vaccine implementation, prevalence is 13.6% (95% CrI: 11.7, 15.3), equal to prevalence in the absence of a vaccine (Figure 1, Table S1). In six of nine epidemic scenarios evaluated, the synergistic effects of risk compensation and viral adaptation led to worse epidemic outcomes than in the absence of a vaccine (Figure 5, Figure S4, S5, S6, Table S2).

**Figure 5.**
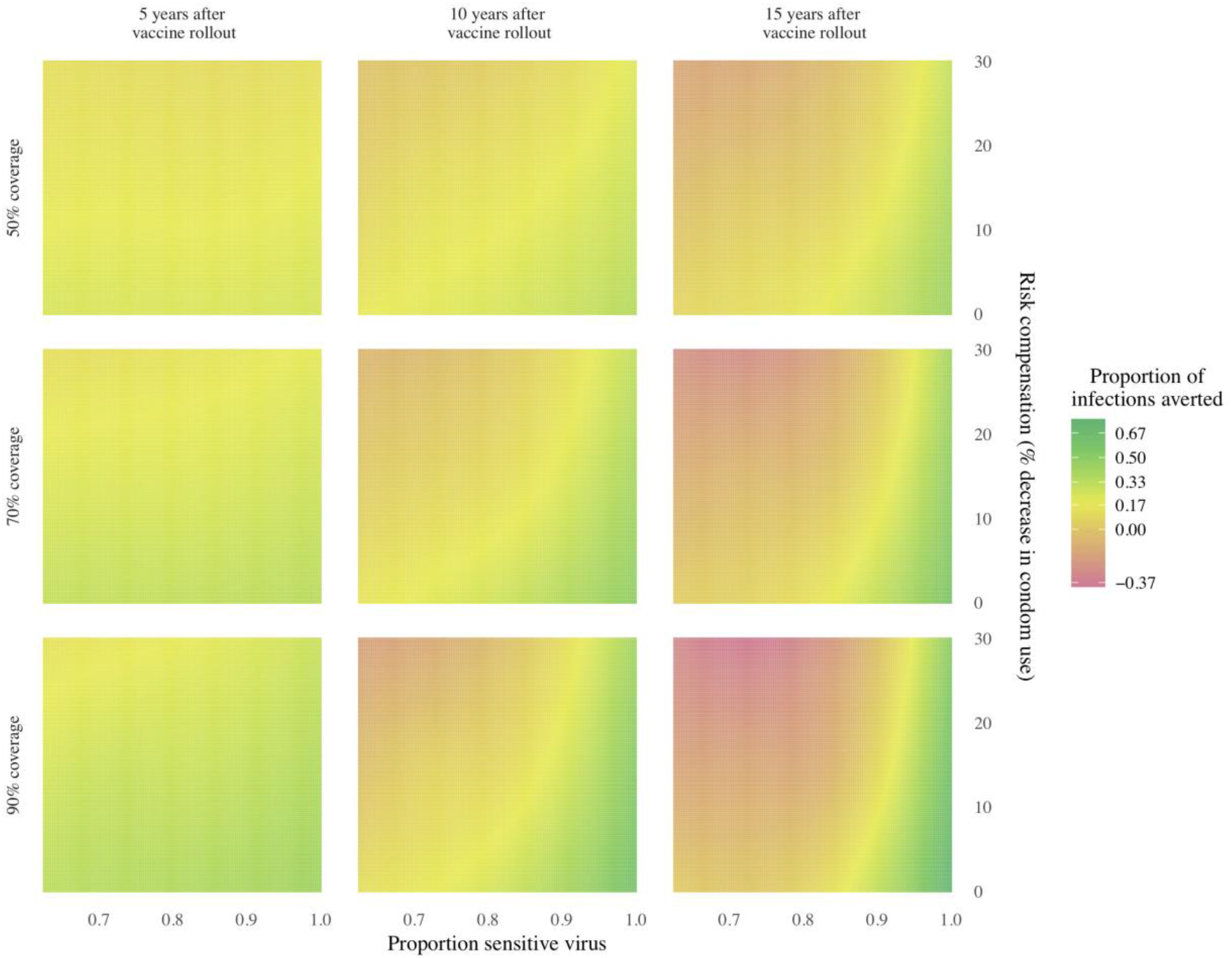
Heatmap of HIV-1 infections averted within 5, 10, and 15 years after vaccine rollout, for multiple combinations of vaccine coverage, risk compensation in vaccinated individuals, and the population frequency of the vaccine resistant virus, relative to a counterfactual model scenario with no vaccine. Perverse vaccine outcomes can occur in scenarios with higher initial frequencies of resistant virus and higher rates of risk compensation (decreased condom use), at all levels of vaccine coverage.

### Cost-effectiveness

We estimated the incremental cost-effectiveness ratio (ICER) per infection averted for a partially effective HIV vaccine in scenarios of viral adaptation and risk compensation. First, in the absence of both vaccine-resistant viral strains and risk compensation, our baseline scenario of an HIV-1 vaccine with 50% efficacy and 70% coverage predicts an ICER of $157 per infection averted (90% CrI: $75, $240) over 15 years of vaccination, compared to scenarios without a vaccine (Table 1, Table S3). Second, while both viral adaptation alone and risk compensation alone increase the ICER per infection averted of an HIV vaccine by approximately 3-fold (Table 1, Figure S7), the concurrent effects of viral adaptation and risk compensation can result in as much as 24-fold reduced cost-effectiveness (e.g. ICER per infection averted of $3,786, for scenarios with 17% resistant virus at vaccine rollout and a 30% decrease in condom use in vaccinated individuals). (We discuss the ICER estimates for DALYs in the Supporting Information.)

**Table 1.**
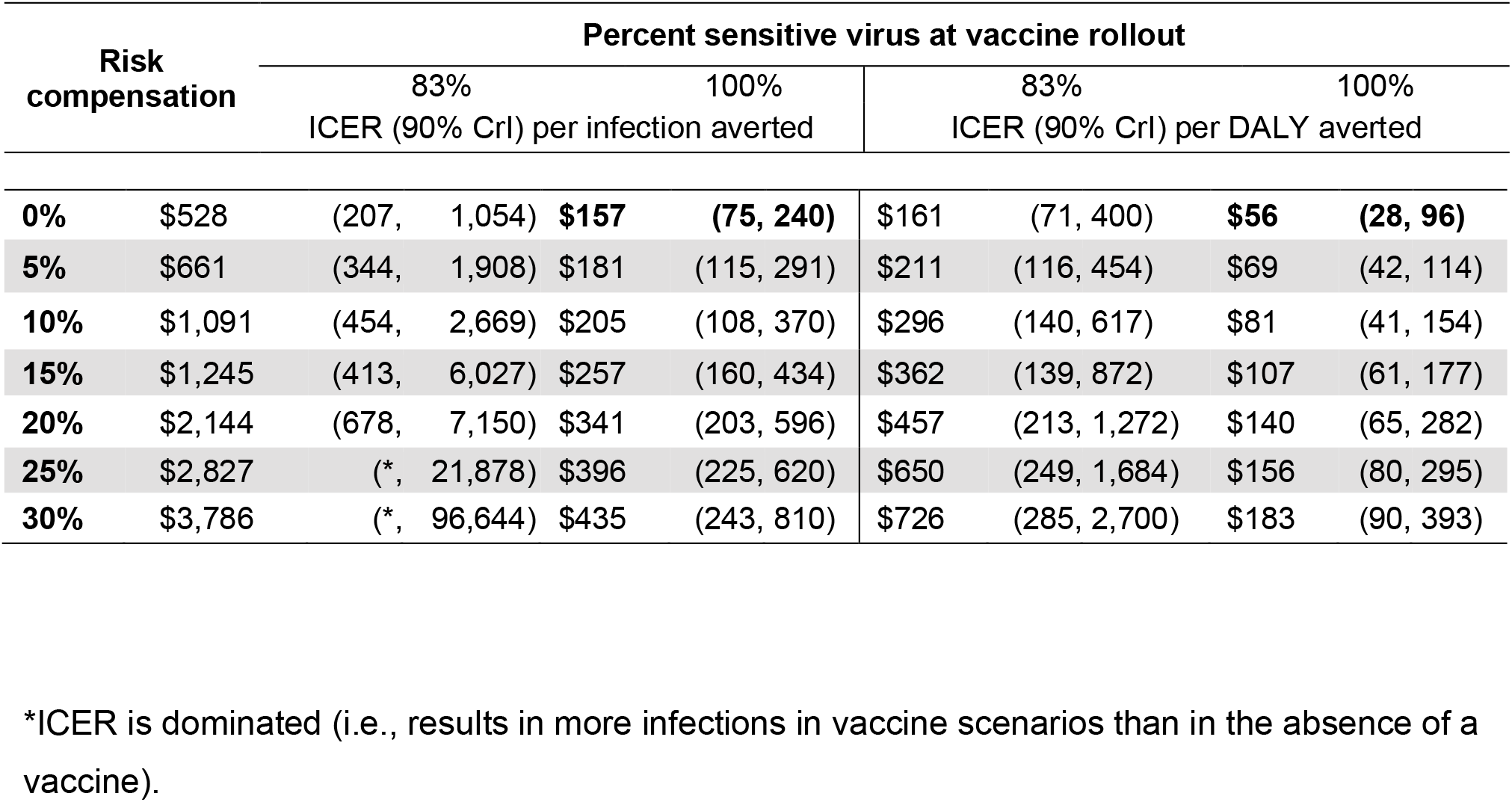
Incremental cost-effectiveness ratio (ICER) per infection averted and per DALY averted over 15 years of continuous vaccine implementation. Costs are in 2017 US dollars. The values in bold text represent the baseline scenario with vaccination, but without risk compensation or resistant virus.

## Discussion

In our simulations of an HIV-1 vaccination program in South Africa, viral adaptation and risk compensation independently and synergistically impacted vaccination campaigns to result in substantially lower epidemic gains than predicted in counterfactual scenarios without these factors. In some scenarios, the combination of viral adaptation and risk compensation resulted in worse epidemic outcomes with a vaccine than without.

These results emphasize the importance of several aspects of vaccine implementation and design. First, viral genomic sieve analyses from all current and future HIV-1 vaccine trials should be conducted to identify viral strains that were differentially impacted by the vaccine. Second, surveillance of HIV-1 diversity in a vaccinated population can inform the risk of viral adaptation and the need for ongoing modification of a vaccine to include inserts (antigens) against alternate viral strains.

Additional HIV-1 vaccine trials of a Mosaic vaccine are currently ongoing in North America, South America, Africa, and Europe (HVTN 705/*Imbokodo* and HVTN 706/*Mosaico*). The Mosaic vaccine is designed to be effective against multiple viral strains, with the expectation that this may also limit rates of viral adaptation. It is possible that concurrent scale-up of additional HIV-1 prevention methods, such as PrEP, may decrease rates of viral evolution (by decreasing total transmissions and rates of viral spread). Additional modeling may inform simultaneous implementation strategies. Finally, communication strategies that balance emphasis of the benefits of an HIV-1 vaccine with an understanding that it is only partially effective may reduce risk compensation among vaccine recipients and the general population.

Viral evolution and risk compensation also independently and synergistically reduce the cost-effectiveness of an HIV-1 vaccine, with their synergistic effects amplifying declines in cost-effectiveness. Counterintuitively, vaccination programs appear cost-effective even in scenarios where long-term epidemic outcomes are worse than in the absence of a vaccine program because cost-effectiveness analyses weight proximal costs and health outcomes more heavily than distal outcomes, emphasizing the need to consider both epidemic outcomes and cost-effectiveness analyses in concert.

There are several limitations in the work presented here. First, we modeled equal fitness in both vaccine-sensitive and vaccine-resistant strains, i.e. sensitive and resistant viruses were not linked to different viral loads or transmissibility in unvaccinated individuals. If, alternately, vaccine resistance was linked with lower intra-host viral loads, we would likely observe slower rates of viral adaptation, as transmission of resistant viruses would be attenuated. Our assumption of equal fitness is supported by the observation that breakthrough infections in RV144 were not associated with decreased viral load or slower rates of HIV disease progression.^2^ Second, our model does not include *de novo* intra-host mutation of vaccine resistance. Previous research, however, has shown that HIV-1 has evolved in response to immune selection pressure;^37-39^ if we included *de novo* mutations in our model to allow escape from the vaccine-elicited immune response, this would likely result in faster population-level viral adaptation. Finally, while there is growing evidence for risk compensation in response to PrEP rollout in high-income settings, evidence of risk compensation in response to biomedical interventions in sub-Saharan Africa is limited to male circumcision in Ugandan and Kenyan populations, rather than the South African population that we have modeled. Additionally, risk compensation may take other forms that are not modeled here, including community-level risk compensation^20,40,41^ and increases in the number of sexual partners. Surveillance of behavioral changes in response to scale up of treatment-as-prevention and PrEP in sub-Saharan Africa might evaluate those forms of risk compensation most relevant to an African context.

In all scenarios, vaccine introduction resulted in epidemic gains in the first five years of rollout. This timeframe suggests there will be an opportunity to monitor and respond to population-level changes in viral strain frequencies and strain-specific incidence. Doing so will require robust surveillance systems with rapid feedback to vaccine design teams, ensuring that modifications to the vaccine can be responsive to epidemic needs. In conclusion, both viral adaptation and risk compensation independently reduce the effectiveness of an HIV-1 vaccine, with their synergistic effects accelerating the timeline of these reductions. Effective implementation of a future HIV-1 vaccine will require robust viral genetic and behavioral surveillance to ensure continued effectiveness.

## Methods

### Mathematical model

We used an individual-based stochastic network model (*EvoNetHIV*, available here: *github*.*com/EvoNetHIV*, and described in detail in the Supplementary Information)^5^ to simulate large-scale vaccine campaigns in populations in which there is viral diversity with respect to vaccine efficacy. The model was calibrated to historical trends in HIV-1 prevalence in South Africa,^42-45^ and included demographics, sexual network structure, sexual behavior, and within-host HIV viral dynamics (Figure S1). We modeled each scenario with an initial population of 30,000, running from 1990 to 2030 (Figure S2). We performed 64 replicate simulations for each scenario. Primary results describe the mean and 95% credible interval of these 64 replicates.

### Vaccine characteristics and scenarios

Because our model was calibrated to the HIV-1 epidemic in South Africa, we modeled an HIV-1 subtype C vaccine with characteristics hypothesized to be similar to the vaccine used in the HVTN 702 trial in South Africa.^46^ The vaccine effect in HVTN 702 was expected to wane over time, similar to waning efficacy observed in RV144,^47^ and HVTN 702 was powered to detect a minimum effect of 50% vaccine efficacy at two years following vaccination. We therefore modeled a partially effective and leaky vaccine, with exponentially decaying (per exposure) vaccine efficacy of the form *VE*_*t*_ = *VE*_0_ − (1 − *e*^−0.000306*e*−4*t*^), with vaccine efficacy duration of five years (VE = vaccine efficacy; *t =* time in days since vaccination). We calculated the initial vaccine efficacy and rate of waning such that the overall population vaccine efficacy at two years in all scenarios, weighted by the proportion of sensitive virus, was equal to 50%. Vaccination in the model begins in year 2018, with user-specified coverage achieved over a three-year period and maintained for the duration of the simulation. Individuals who receive the vaccine and remain HIV-1-negative for the duration of vaccine efficacy are revaccinated at five years. All simulations model 15 years of continuous vaccination coverage.

We modeled viral diversity with respect to vaccine efficacy as two genotypes (i.e. one locus with two alleles) that determine whether a vaccinated individual is conferred a degree of protection or no protection against HIV-1 infection, given a sexual contact with an HIV-infected individual. In our primary analysis we initialized standing viral diversity at vaccine rollout to include 17% vaccine-resistant virus (83% vaccine-sensitive) and compared this scenario to a counterfactual in which 100% of virus is vaccine sensitive. We included additional scenarios in which 29% and 37% of virus is vaccine-resistant at rollout as sensitivity analyses. We also evaluated variation in vaccine coverage, assessing 70% coverage in our primary analyses and 50% and 90% coverage in sensitivity analyses.

Risk compensation in response to biomedical interventions has been observed to be as high as a 75% increase in high-risk sexual behavior in high-income settings,^16,48^ though has been lower where observed in sub-Saharan Africa, ranging from 12%^24^ to 18%.^22^ Evaluations of risk compensation have additionally focused primarily on condom use, and, though less frequently assessed, have rarely found changes in the number of sexual partners.^17,18,22^ We therefore model risk compensation as a relative reduction in the per-act probability of condom use among vaccinated individuals, evaluating the range of 5%-30% reduction. Baseline per-act probabilities of condom use increase over time according to nationally representative surveys in South Africa from 2002-2012.^42-45^

### Estimation of vaccine cost-effectiveness

We calculated the incremental cost-effectiveness ratio (ICER) for the vaccine program, using two health outcomes: infections averted and disability-adjusted life years (DALY) averted. To estimate DALYs, we tracked the number of person-years spent in the following categories: 1) HIV-infected with CD4+ T cell count >350; 2) HIV-infected with CD4 count between 200 and 350; 3) HIV-infected with CD4 <200; and 4) HIV-infected and on ART. The number of person-years in each category was then multiplied by the corresponding disability weight to calculate category-specific DALYs.^49^ These, along with years of life lost due to premature AIDS death, were then summed to total DALYs and used in comparisons across scenarios. We used sex-specific life expectancy estimates for the year 2017 in estimation of DALYs.^50^ Estimates of cost-effectiveness include the incremental cost of the vaccination program, assuming a cost of $60 per vaccine series, following other models of an HIV-1 vaccine in middle-income countries,^34^ as well as HIV-1 care and treatment costs averted by the vaccine program.^51^ Both costs and health outcomes are discounted at an annual rate of 3%.^52^

## Data Availability

All model code is freely available on github.com/EvoNetHIV.

## Acknowledgements

This work was supported by the U.S. National Institutes of Health grants R01-AI108490 (SMG, JTH, JEM), R01-AI150467 (JTH), R21-HD075662 (SMG), R01-HD068395 (provided support to SMG), and the University of Washington student technology fee (https://uwstf.org) (support for the Hyak super-computer system used in this research). The funders had no role in study design, data collection and analysis, decision to publish, or preparation of the manuscript.

## Author information

### Contributions

K.P. and J.T.H. conceived and designed the study and drafted the manuscript. K.P. performed the model analyses. K.P., J.E.M., S.M.G., J.T.M., M.C.R., N.A., G.S.G., and J.T.H. developed the epidemiological and evolutionary model. K.P. and R.V.B. designed the cost effectiveness analyses. All authors reviewed and interpreted the results and contributed to writing the final manuscript.

