## Supplementary Info for "Risk compensation after HIV-1 vaccination may accelerate viral adaptation and reduce cost-effectiveness: a modeling study"

*^1^Department of Epidemiology, ^2^Department of Microbiology, ^3^Center for Studies in Demography and Ecology, ^4^Department of Anthropology, ^5^Department of Global Health, ^6^Department of Biomedical Inf. and Medical Education, ^7^Department of Health Services, ^8^Department of Medicine, ^9^Center for Emerging & Re-Emerging Infectious Diseases, ^10^International Clinical Research Center, University of Washington, Seattle, WA*

**Model description**

The simulations described in this paper use the *EvoNetHIV* epidemic and evolutionary model framework described in detail in the Methods and Supplementary Material of previous publications.^1-4^ All model code was written in R and C++ and constructed using the *EpiModel* package API^5^ and is available at <https://github.com/EvoNetHIV/>.

*EvoNetHIV* is a stochastic individual-based model that tracks inter-host and intra-host viral dynamics, relationship dynamics, and demographic dynamics at daily time-steps. All individuals were assigned demographic, clinical, and behavioral traits. Each simulation was initiated with HIV-1-negative and -positive individuals. The dynamics of sexual relationships were governed by network models based in the separable-temporal exponential-family random graph model (STERGM) framework.^6^ Partnerships formed and dissolved to match target network parameters. Dyads formed and dissolved partnerships to match target network parameters, including gender-asymmetric age-matching such that, on average, males are three years older than their female partners.^3^

Within ongoing partnerships, we modeled individual coital acts, as well as decisions about condom use per act. For each serodiscordant coital act that occurred, the probability of transmission from an HIV-1-positive person to an HIV-1-negative person was based primarily on instantaneous viral load at the time step of the contact, but also on disease stage (above viral load), age, sex, condom use, and antiretroviral use; this is described by a complementary-log log link function that follows from Hughes et al.^7^

HIV-1-infected individuals possess viral characteristics such as the “set point viral load” (SPVL) and a viral load at each time step over the course of infection. The individual SPVL is determined by both viral and environmental (stochastic) factors, with the viral component being heritable across transmissions. For all individuals, upon HIV-1 infection with an initial viral population size, viral load increases exponentially until a peak viremia at the midpoint of the length of acute infection. Viral load then declines exponentially at an individual-specific decay rate until it reaches SPVL. After reaching set point, the viral load increases in a log linear manner until the onset of AIDS. Viral load upon onset of AIDS is defined as the same for all individuals and is independent of SPVL.

To model the relationship between SPVL and disease progression, the model relates individual SPVL to the starting CD4 count category (individual CD4 count immediately after infection) and to disease progression based on waiting times in four CD4 count categories: CD4≥500; 500>CD4≥350; 350>CD4≥200; CD4<200 (AIDS).^8^

**Model calibration**

We calibrated the simulated heterosexual HIV epidemic to reproduce the observed age- and sex-specific prevalence trajectories reported in nationally representative South African surveys from 2002 to 2012.^9-12^ We chose to calibrate the model to the South African epidemic for the following reasons: 1) because South Africa has one of the largest HIV epidemics; 2) because the HIV vaccine trial HVTN 702 was located in South Africa, and is based upon the RV144 trial design and results; and 3) so that the simulated epidemic output could be comparable to the 12 HIV epidemic models described in Eaton et al.^13^ Using an approach common among those models, we first set empirical parameter values (i.e., viral load trajectories, CD4 progression rates, set point viral load distribution), then subsequently calibrated the assumption-based parameter values to fit expected epidemic dynamics.

We calibrated the model using an approximate Bayesian computation model selection procedure using the *easyABC* R package. Values for seven per-act infectivity and relationship duration and concurrency parameters were selected via an ABC Sequential Monte Carlo sampling process; from among 500 simulations using these fitted values, we selected the simulation that best fit observed prevalence data by least-squares, shown in Figure S1. We use this single simulation for the first 28 years of each modeling scenario, representing the years 1990-2018. For our final analyses concerning viral adaptation, risk compensation, and vaccine impact, we note that the calibration fit to South Africa is not in itself critical, as our experimental procedure compares output from identically calibrated model simulations that vary only in vaccine-related parameters (which are not included in the original model calibration).

**Condom use**

Condom use is modeled as a Hill function to fit population-based estimates of condom use over time (Figure S9).^9-12^ We assume that condom use in the early 1990’s was negligible, with a sigmoidal scale-up to fit observed data of the form:

$$condom probability = 0.04+ \frac{0.50 \times t^{4.1}}{{12}^{4.1}+t^{4.1}}$$

where *t* indicates the model time step. Condom use reduces the risk of HIV acquisition by 78%.^7^

**Risk compensation**

We modeled risk compensation as a relative reduction in the per-act probability of condom use among vaccinated individuals, evaluating the range of 5%-30% reduction.

**Vaccination**

We modeled our vaccine effect after that effect observed in the HIV vaccine trial RV144, which is described by the vaccine efficacy waning function in Hankins, et al., 2011^14^:

$${VE}_{t}={VE}_{0}-(1-e^{-0.0003056761t})$$

VE_0_ is vaccine efficacy on day 1 of vaccination and *t* is time in days since vaccination. The formula above produces VE at year two that, when considering the proportion of resistant virus, would result in observed 50% VE in the trial context in which, due to a small proportion of the population being enrolled in the trial, viral adaptation should not be observed. Note that in our model, viral adaptation will be observed, as the entire population is eligible for vaccination, and so VE at year 2 will be lower than 50%. Vaccinated individuals who remain HIV-negative for the duration of the vaccine’s five-year efficacy period will be re-vaccinated once five years have elapsed since their previous vaccination. Figure S10 shows intra-individual vaccine waning and boosting over the course of 15 years for each of the experimental scenario VEs.

**Antiretroviral treatment**

We modeled the use of antiretroviral treatment (ART). ART is distributed beginning in 2001 per CD4-based national guidelines, which are modified in 2011, 2014, and 2016 to reflect decreasingly restrictive eligibility criteria. ART coverage among eligible individuals increases according to estimates of observed ART coverage (Table S5).^15,16^ ART coverage among all eligible individuals is scaled to reflect observed age- and sex-based disparities in coverage (Table S6).^12^

**Results: Cost-effectiveness per DALY**

We estimated the incremental cost-effectiveness ratio (ICER) per disability adjusted life year (DALY) for a partially effective HIV vaccine in scenarios of viral adaptation and risk compensation (Figure S8). First, in the absence of both vaccine resistant viral strains and risk compensation, our baseline scenario of an HIV-1 vaccine with 50% efficacy and 70% coverage predicted an ICER of $56 per DALY (90% CrI: $28, $96) over 15 years of vaccination, compared to scenarios without a vaccine (Table 1). Second, while both viral adaptation alone and risk compensation alone increased the ICER per DALY of an HIV vaccine by approximately 3-fold (i.e. $161 per DALY), the concurrent effects of viral adaptation and risk compensation resulted in cost increases of as much as 13-fold. (e.g. ICER per DALY of $726, for scenarios with 17% resistant virus at vaccine rollout and a 30% decrease in condom use in vaccinated individuals).

16	UNAIDS. UNAIDS Data 2020. (Geneva, 2020).

**Table S5**. Coverage of antiretroviral treatment (ART) among treatment-eligible individuals in South Africa.

| **Year** | **ART coverage** | **Source** |
| --- | --- | --- |
| 2001 | 0.010 | Adam, 2009 |
| 2002 | 0.021 | Adam, 2009 |
| 2003 | 0.030 | Adam, 2009 |
| 2004 | 0.049 | Adam, 2009 |
| 2005 | 0.100 | Adam, 2009 |
| 2006 | 0.191 | Adam, 2009 |
| 2007 | 0.283 | Adam, 2009 |
| 2008 | 0.402 | Adam, 2009 |
| 2016 | 0.560 | UNAIDS, 2020 |

**Table S6**. Age- and sex-specific antiretroviral coverage adjustment scalars.

| Age | Sex | |
| --- | --- | --- |
|  | Female | Male |
| 15-24 | 0.569 | 0.421 |
| 25-49 | 1.240 | 0.919 |
