## Supplementary Tables for "Risk compensation after HIV-1 vaccination may accelerate viral adaptation and reduce cost-effectiveness: a modeling study"

**Table S1**. HIV prevalence following 15 years of continuous vaccine coverage, mean (95% credible interval). Final prevalence in a counterfactual scenario in which no vaccine is implemented is **13.6% (95% CrI: 12.5, 14.9)**. The value in bold text represents the baseline scenario with vaccination, but without risk compensation or resistant virus.

| Percent sensitive virus at rollout | Coverage | Percent reduction in condom use | | | | | | |
| --- | --- | --- | --- | --- | --- | --- | --- | --- |
|  |  | 0% | 5% | 10% | 15% | 20% | 25% | 30% |
| 62.5% sensitive virus | 50% | 11.5 (9.9, 12.7) | 12.0 (10.2, 13.3) | 12.4 (11.2, 13.7) | 12.8 (11.2, 13.7) | 13.2 (11.8, 14.2) | 13.5 (11.6, 15.3) | 14.1 (12.1, 15.3) |
|  | 70% | 11.6 (10.2, 12.9) | 12.1 (10.6, 13.4) | 12.7 (11.3, 13.9) | 13.3 (11.7, 14.6) | 13.5 (11.4, 14.7) | 14.1 (12.5, 15.4) | 14.9 (13.3, 16.2) |
|  | 90% | 11.8 (10.2, 13.1) | 12.3 (11.0, 13.6) | 13.1 (11.5, 14.5) | 13.6 (12.1, 14.7) | 14.4 (12.8, 15.6) | 15.0 (13.3, 16.2) | 15.4 (14.1, 16.7) |
| 71.4% sensitive virus | 50% | 10.9 (9.4, 12.3) | 11.4 (9.1, 13.0) | 11.7 (10.5, 13.2) | 12.3 (10.0, 14.0) | 12.6 (10.8, 14.1) | 13.2 (11.4, 14.7) | 13.3 (11.2, 14.6) |
|  | 70% | 10.9 (9.6, 12.2) | 11.5 (9.6, 13.3) | 12.0 (9.6, 13.4) | 12.7 (10.8, 14.2) | 13.4 (11.7, 14.6) | 13.5 (11.4, 14.8) | 14.2 (12.6, 15.7) |
|  | 90% | 11.1 (9.5, 12.9) | 11.8 (10.6, 12.8) | 12.3 (10.3, 13.5) | 13.0 (11.1, 14.7) | 13.6 (12.1, 14.8) | 14.3 (12.4, 15.5) | 15.0 (12.7, 16.5) |
| 83.3% sensitive virus | 50% | 10.9 (9.1, 12.4) | 11.0 (9.3, 12.3) | 11.4 (9.5, 13.0) | 11.9 (10.2, 13.3) | 12.3 (10.3, 14.0) | 12.7 (11.4, 14.2) | 13.2 (11.4, 15.0) |
|  | 70% | 10.4 (8.3, 11.8) | 11.0 (9.4, 12.7) | 11.6 (9.9, 12.9) | 11.9 (9.7, 13.7) | 12.7 (10.9, 14.0) | 13.2 (11.1, 14.9) | 13.6 (11.7, 15.3) |
|  | 90% | 10.7 (8.8, 12.5) | 11.1 (9.3, 12.6) | 12.0 (10.1, 13.4) | 12.5 (9.9, 14.2) | 13.0 (11.2, 14.6) | 13.6 (11.4, 15.7) | 14.3 (12.1, 15.9) |
| 100% sensitive virus | 50% | 9.0 (7.9, 9.7) | 9.2 (8.3, 10.0) | 9.5 (8.6, 10.6) | 9.7 (8.8, 10.5) | 10.0 (9.0, 10.9) | 10.2 (9.1, 11.2) | 10.6 (9.4, 11.4) |
|  | 70% | **7.8 (7.0, 8.4)** | 8.0 (7.3, 8.7) | 8.2 (7.4, 8.9) | 8.5 (7.6, 9.2) | 8.9 (8.2, 9.7) | 9.0 (8.3, 9.8) | 9.3 (8.6, 10.2) |
|  | 90% | 6.7 (6.0, 7.2) | 7.0 (6.3, 7.4) | 7.2 (6.6, 7.8) | 7.5 (6.8, 8.2) | 7.8 (7.2, 8.5) | 8.0 (7.1, 8.6) | 8.3 (7.6, 9.0) |

**Table S2**. Percent infections averted following 15 years of continuous vaccine implementation, mean (95% credible interval), compared to a counterfactual scenario in which no vaccine is implemented. The value in bold text represents the baseline scenario with vaccination, but without risk compensation or resistant virus.

| Percent sensitive virus at rollout | Coverage | Percent reduction in condom use | | | | | | |
| --- | --- | --- | --- | --- | --- | --- | --- | --- |
|  |  | 0% | 5% | 10% | 15% | 20% | 25% | 30% |
| 62.5% sensitive virus | 50% | 7.9 (-11.1, 22.9) | 3.6 (-12.7, 19.0) | -0.3 (-19.2, 19.2) | -4.5 (-27.0, 14.2) | -10.8 (-26.0, 6.2) | -11.7 (-38.4, 8.4) | -18.1 (-36.0, 2.3) |
|  | 70% | 4.4 (-16.9, 23.8) | -1.2 (-14.5, 16.3) | -8.2 (-27.2, 15.1) | -13.5 (-30.6, 3.0) | -14.5 (-32.1, 1.6) | -20.5 (-36.9, 2.5) | -27.5 (-56.5, -11.2) |
|  | 90% | 2.1 (-17.0, 23.3) | -3.4 (-23.2, 14.6) | -11.6 (-31.6, 8.7) | -17.8 (-34.7, 0.2) | -21.8 (-42.5, -1.1) | -26.5 (-43.9, -5.2) | -31.6 (-49.0, -7.4) |
| 71.4% sensitive virus | 50% | 13.9 (-7.8, 32.4) | 11.8 (-11.9, 30.0) | 5.7 (-13.6, 23.2) | 0.9 (-24.2, 25.3) | -3.6 (-22.4, 19.0) | -11.1 (-37.2, 9.7) | -8.7 (-25.9, 10.4) |
|  | 70% | 10.7 (-14.0, 29.2) | 3.5 (-15.6, 24.7) | 0.9 (-21.2, 28.8) | -6.3 (-27.6, 14.8) | -14.1 (-35.0, 1.5) | -16.5 (-34.2, -0.4) | -23.6 (-41.2, -3.3) |
|  | 90% | 6.2 (-19.1, 25.0) | -2.0 (-20.8, 16.0) | -6.9 (-27.2, 17.4) | -15.1 (-31.4, 3.3) | -21.5 (-44.4, 2.8) | -26.5 (-45.3, -3.5) | -31.8 (-51.1, -14.8) |
| 83.3% sensitive virus | 50% | 17.3 (-8.7, 40.8) | 15.5 (-8.7, 35.9) | 10.2 (-13.6, 37.9) | 4.3 (-17.4, 24.8) | 1.2 (-23.6, 22.2) | -3.6 (-26.4, 19.4) | -8.8 (-28.4, 10.1) |
|  | 70% | 16.8 (-4.4, 41.5) | 10.0 (-12.1, 28.9) | 2.5 (-18.7, 24.2) | -0.3 (-28.6, 24.4) | -8.5 (-32.9, 8.8) | -15.6 (-40.5, 6.4) | -19.9 (-41.0, 0.3) |
|  | 90% | 9.7 (-9.1, 35.3) | 5.3 (-13.6, 24.4) | -6.2 (-25.9, 11.7) | -10.5 (-30.8, 19.5) | -16.8 (-38.7, 4.5) | -20.6 (-38.0, -1.8) | -29.3 (-46.6, -1.9) |
| 100% sensitive virus | 50% | 44.0 (31.6, 56.5) | 42.3 (29.9, 53.8) | 40.2 (27.1, 52.1) | 36.8 (23.5, 46.3) | 33.3 (19.6, 46.7) | 32.5 (17.1, 42.9) | 29.3 (16.8, 42.9) |
|  | 70% | **56.8 (41.4, 69.0)** | 55.1 (45.3, 66.4) | 51.8 (38.3, 63.1) | 49.2 (30.5, 62.6) | 44.9 (34.8, 53.8) | 43.5 (34.3, 54.8) | 41.6 (29.6, 51.6) |
|  | 90% | 73.9 (68.2, 81.6) | 69.6 (56.9, 79.0) | 66.5 (55.9, 76.3) | 62.1 (47.3, 72.9) | 57.5 (43.2, 68.8) | 54.4 (41.9, 65.0) | 50.5 (39.8, 59.0) |

**Table S3**. Incremental cost-effectiveness ratio (ICER) per infection averted over 15 years of continuous vaccine implementation, mean (90% credible interval). Costs are in 2017 US dollars. The value in bold text represents the baseline scenario with vaccination, but without risk compensation or resistant virus.

| Percent sensitive virus at rollout | Coverage | Percent reduction in condom use | | | | | | |
| --- | --- | --- | --- | --- | --- | --- | --- | --- |
|  |  | 0% | 5% | 10% | 15% | 20% | 25% | 30% |
| 62.5% sensitive virus | 50% | 456 (106, 1,480) | 672 (183, 1,894) | 1,172 (403, 4,253) | 1,647 (566, 6,377) | 2,591 (433, 19,476) | 2,125 (*, 17,354) | 2,009 (*, 21,289) |
|  | 70% | 960 (466, 2,227) | 1,236 (559, 3,366) | 1,864 (792, 4,951) | 3,106 (1,046, 9,913) | 3,279 (*, 39,699) | 5,298 (*, 41,040) | * (*, 49,790) |
|  | 90% | 1,480 (891, 2,711) | 1,958 (1,097, 3,929) | 2,957 (1,260, 8,039) | 4,020 (1,558, 13,039) | 7,327 (*, 61,462) | * (*, 54,217) | * (*, 31,888) |
| 71.4% sensitive virus | 50% | 259 (*, 955) | 542 (75, 1,550) | 567 (187, 2,005) | 1,018 (197, 3,939) | 1,570 (310, 10,088) | 1,650 (*, 13,784) | 1,977 (*, 13,001) |
|  | 70% | 666 (303, 1,274) | 906 (435, 3,080) | 1,260 (454, 3,625) | 1,900 (693, 7,481) | 2,784 (*, 11,762) | 3,387 (*, 25,508) | 2,801 (*, 14,245) |
|  | 90% | 1,006 (568, 2,038) | 1,496 (824, 2,486) | 1,876 (1,077, 4,202) | 2,964 (1,196, 8,489) | 3,634 (1,176, 15,912) | 6,398 (*, 41,812) | * (*, 42,118) |
| 83.3% sensitive virus | 50% | 332 (40, 1,251) | 407 (43, 1,164) | 598 (87, 1,419) | 779 (198, 3,819) | 1,064 (209, 4,893) | 1,720 (610, 7,818) | 2,256 (*, 23,068) |
|  | 70% | 528 (207, 1,054) | 661 (344, 1,908) | 1,091 (454, 2,669) | 1,245 (413, 6,027) | 2,144 (678, 7,150) | 2,827 (*, 21,878) | 3,786 (*, 96,644) |
|  | 90% | 893 (503, 1,615) | 1,095 (607, 2,081) | 1,493 (764, 4,100) | 1,986 (986, 7,582) | 2,954 (1,211, 20,691) | 3,283 (*, 19,337) | 4,494 (*, 29,647) |
| 100% sensitive virus | 50% | 19 (†, 170) | 53 (†, 227) | 114 (†, 335) | 177 (†, 428) | 237 (12, 591) | 346 (68, 915) | 494 (88, 1,137) |
|  | 70% | **157 (75, 240)** | 181 (115, 291) | 205 (108, 370) | 257 (160, 434) | 341 (203, 596) | 396 (225, 620) | 435 (243, 810) |
|  | 90% | 260 (205, 314) | 290 (208, 377) | 319 (260, 429) | 362 (277, 465) | 404 (315, 566) | 472 (360, 627) | 511 (389, 660) |

* ICER is dominated (i.e., results in more infections in vaccine scenarios than in the absence of a vaccine).

† Cost-saving

**Table S4**. Incremental cost-effectiveness ratio (ICER) per disability-adjusted life-year averted over 15 years of continuous vaccine implementation, mean (90% credible interval). Costs are in 2017 US dollars. The value in bold text represents the baseline scenario with vaccination, but without risk compensation or resistant virus.

| Percent sensitive virus at rollout | Coverage | Percent reduction in condom use | | | | | | |
| --- | --- | --- | --- | --- | --- | --- | --- | --- |
|  |  | 0% | 5% | 10% | 15% | 20% | 25% | 30% |
| 62.5% sensitive virus | 50% | 140 (32, 357) | 230 (66, 507) | 261 (114, 915) | 376 (125, 1,116) | 479 (186, 3,763) | 561 (217, 2,599) | 716 (183, 4,118) |
|  | 70% | 269 (135, 587) | 311 (149, 779) | 437 (195, 733) | 549 (272, 1,131) | 567 (187, 1,270) | 838 (404, 3,664) | 1,104 (*, 7,218) |
|  | 90% | 341 (232, 573) | 433 (276, 826) | 525 (296, 899) | 598 (398, 1,685) | 1,022 (530, 2,514) | 1,288 (593, 16,987) | 1,666 (448, 6,533) |
| 71.4% sensitive virus | 50% | 95 (†, 283) | 171 (27, 385) | 187 (64, 588) | 303 (77, 702) | 399 (97, 934) | 473 (146, 2,374) | 485 (178, 2,309) |
|  | 70% | 198 (88, 374) | 242 (127, 653) | 330 (143, 927) | 447 (172, 924) | 560 (297, 1,509) | 602 (266, 1,760) | 907 (341, 3,537) |
|  | 90% | 258 (160, 482) | 325 (206, 571) | 421 (273, 798) | 527 (290, 897) | 610 (315, 1,202) | 859 (472, 2,500) | 1,385 (464, 4,419) |
| 83.3% sensitive virus | 50% | 110 (14, 336) | 151 (15, 358) | 190 (34, 435) | 251 (80, 735) | 371 (107, 1,306) | 469 (200, 1,396) | 634 (105, 2,269) |
|  | 70% | 161 (71, 400) | 211 (116, 454) | 296 (140, 617) | 362 (139, 872) | 457 (213, 1,272) | 650 (249, 1,684) | 726 (285, 2,700) |
|  | 90% | 253 (153, 442) | 297 (175, 477) | 378 (204, 715) | 442 (257, 873) | 576 (308, 1,448) | 805 (334, 2,260) | 998 (470, 3,283) |
| 100% sensitive virus | 50% | 7 (†, 82) | 23 (†, 110) | 52 (†, 164) | 73 (†, 186) | 99 (5, 271) | 144 (29, 450) | 198 (34, 534) |
|  | 70% | **56 (28, 96)** | 69 (42, 114) | 81 (41, 154) | 107 (61, 177) | 140 (65, 282) | 156 (80, 295) | 183 (90, 393) |
|  | 90% | 93 (69, 118) | 103 (71, 139) | 116 (89, 166) | 129 (99, 178) | 150 (111, 221) | 175 (131, 269) | 188 (131, 281) |

* ICER is dominated (i.e., results in more disability-adjusted life-years in vaccine scenarios than in the absence of a vaccine).

† Cost-saving
