## Supplementary Figures for "Risk compensation after HIV-1 vaccination may accelerate viral adaptation and reduce cost-effectiveness: a modeling study"

**Figure S1.** Model calibration to observed age- and sex-specific HIV prevalence reported in nationally representative South Africa surveys from 2002 to 2012 in an approximate Bayesian computation model selection procedure. The single best-fitting calibrated simulation was applied to the years 1990-2018 for each modeling scenario.

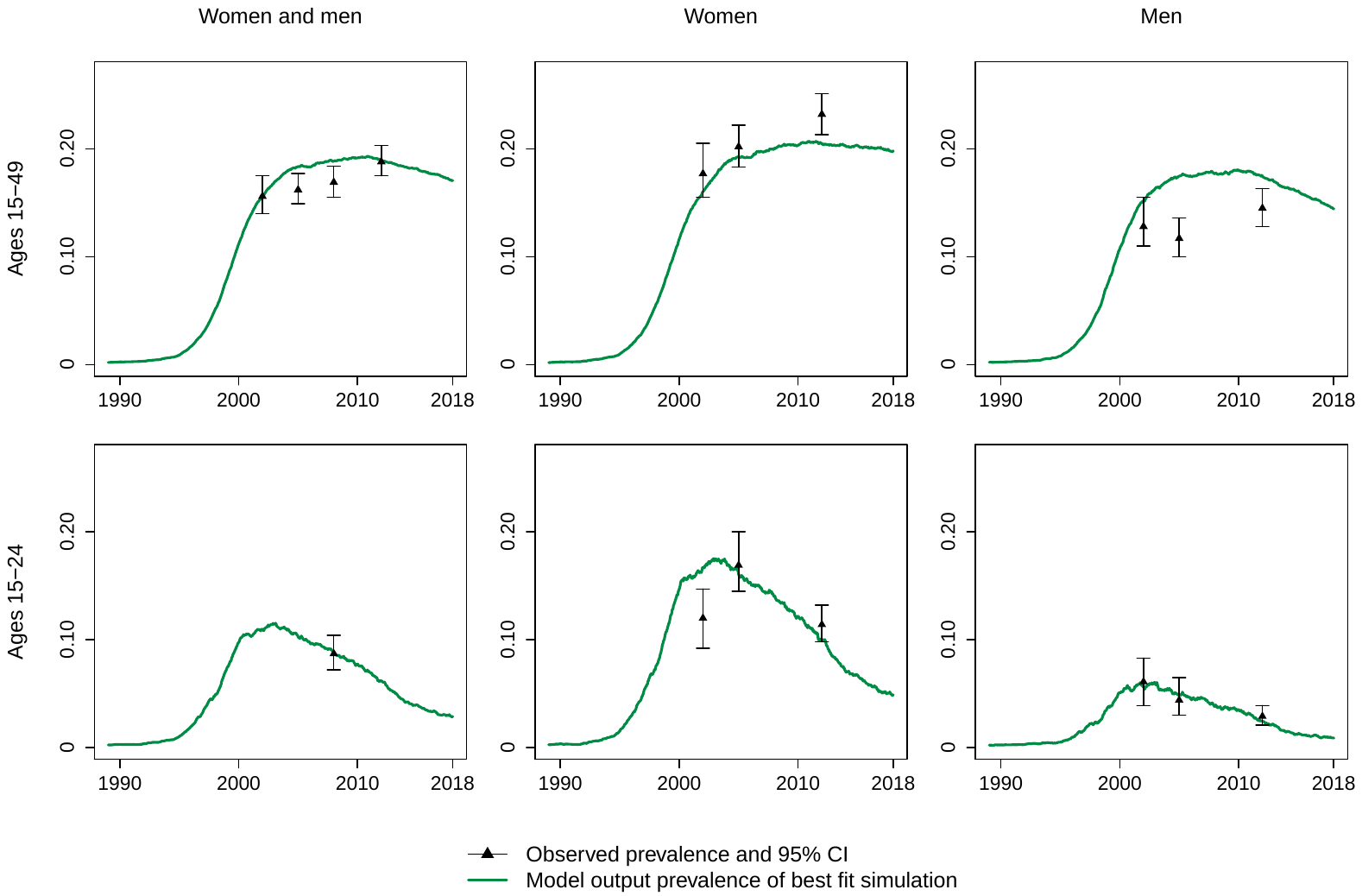

**Figure S2.** Simulated HIV incidence plots from 1990 to 2035 for the model calibration used in these analyses. Overall incidence is shown in black, incidence of vaccine-sensitive viral infections is in pink, and incidence of vaccine-resistant viral infections is in green. The **top** plot depicts a model scenario with no vaccine and no transmission advantage to the vaccine resistant virus. The **bottom** plot depicts a model scenario with sensitive and resistant viruses, a transmission advantage to the resistant virus, and behavioral risk compensation. The vertical dashed line marks the year of vaccine rollout in 2018 *only for* the model scenarios with vaccination (the bottom plot in the Figure). All scenarios had the same epidemic trajectory prior to 2018; after 2018 each scenario was repeated with 64 replicates and the plots show the mean and 95% credible intervals for each scenario.

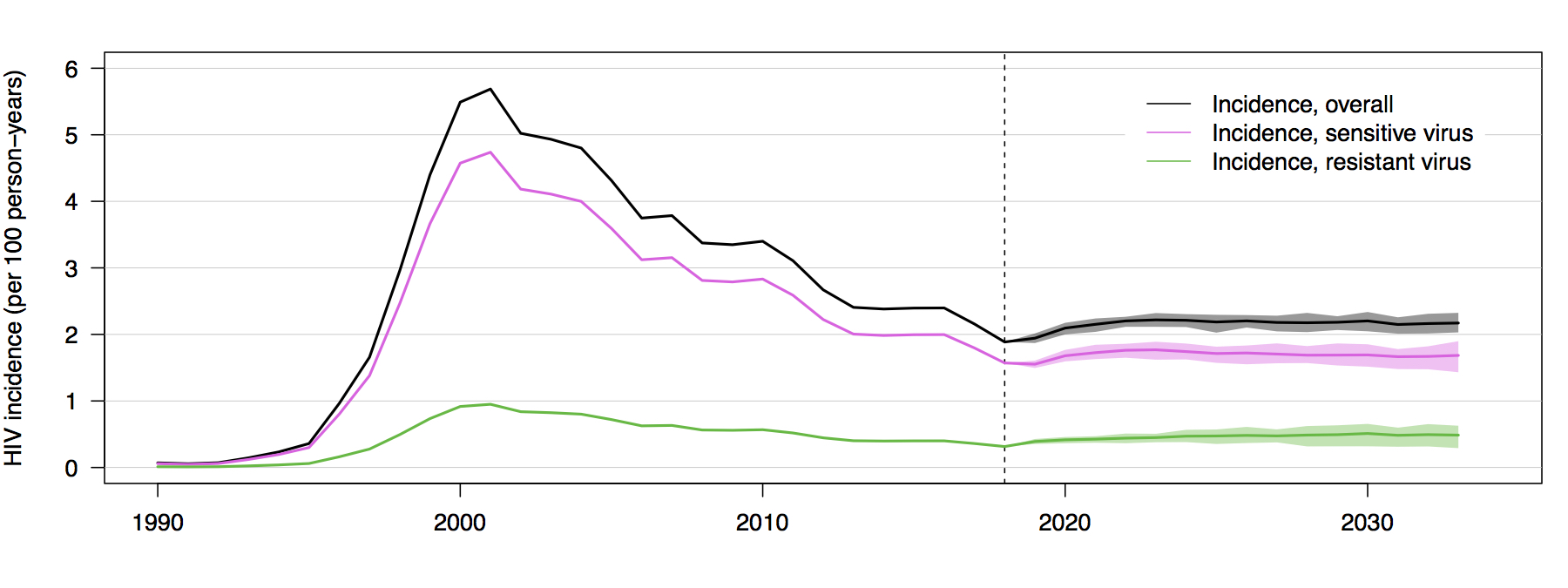

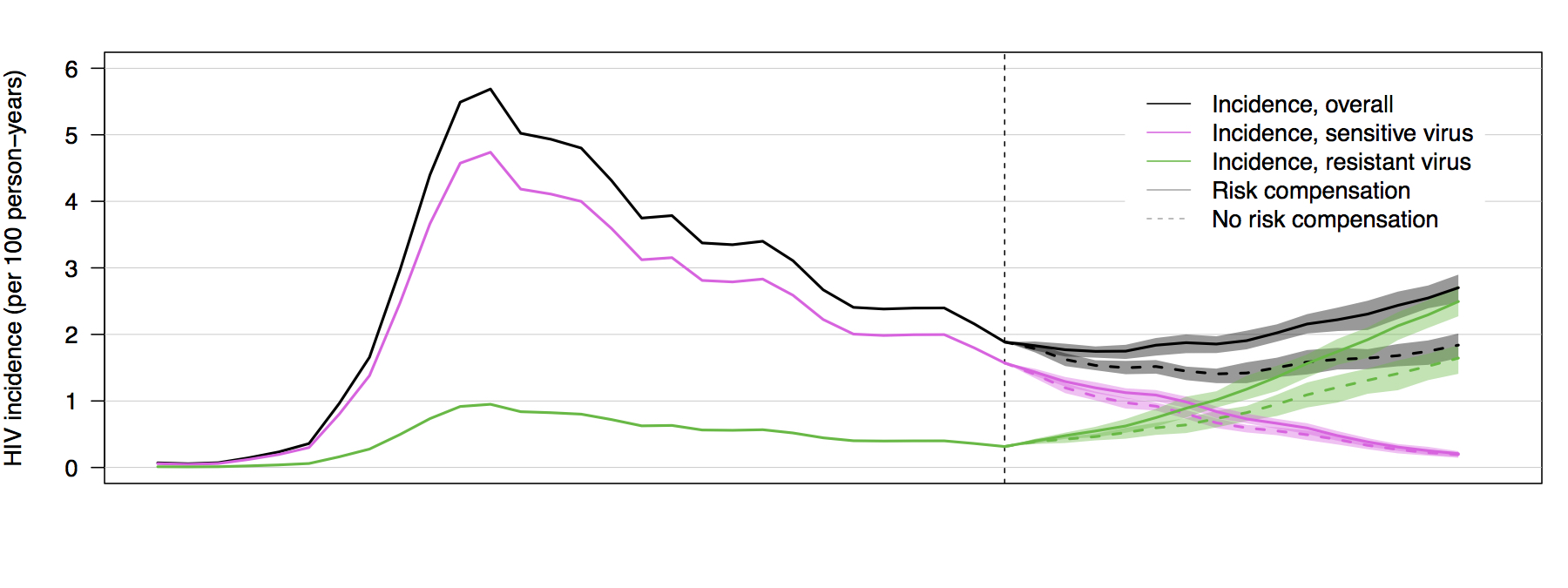

**Figure S3.** Frequencies of HIV lineages that are either sensitive or resistant to vaccine induced immune responses, following introduction of a partially effective vaccine (here shown as years from vaccine rollout). Each plot shows the mean and 95% credible interval of 64 replicates of each scenario. The scenarios vary in the proportion of vaccine sensitive virus at vaccine rollout (62.5%, 71.4%, 83.3%), variation in vaccine population coverage (50%, 70%, 90%), and, within each plot, the presence or absence of risk compensation (0% vs 30% reduction in condom use among vaccinated individuals).

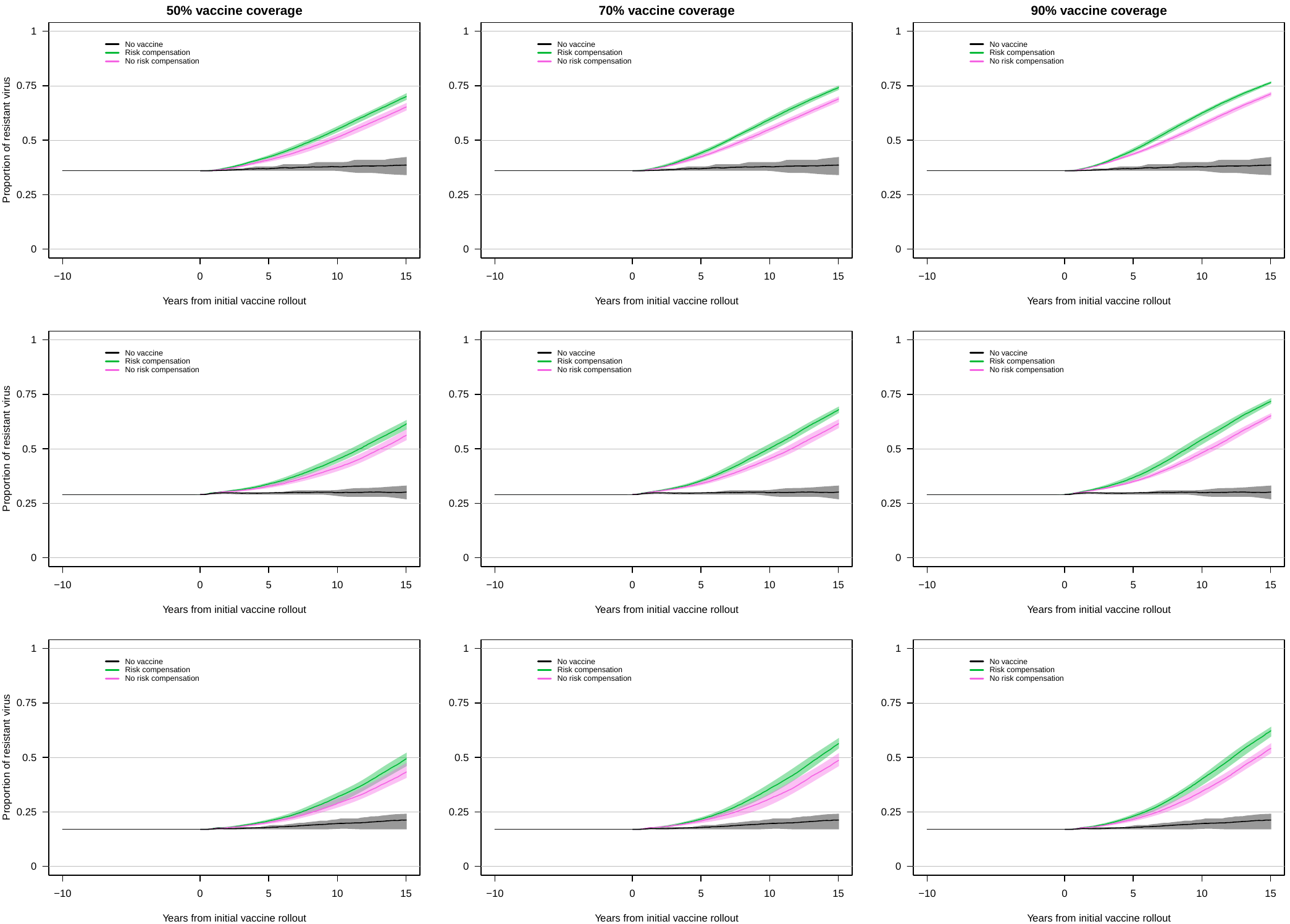

**Figure S4**. HIV-1 prevalence following introduction of a partially effective vaccine in the year 2018. Each figure shows the mean and 95% credible interval of 64 replicates of each scenario. Scenarios include variation in the proportion of vaccine-sensitive virus at vaccine rollout (62.5%, 71.4%, and 83.3% compared to 100%) and variation in the reduction in condom use among vaccinated individuals (from 0% to 30% reduction).

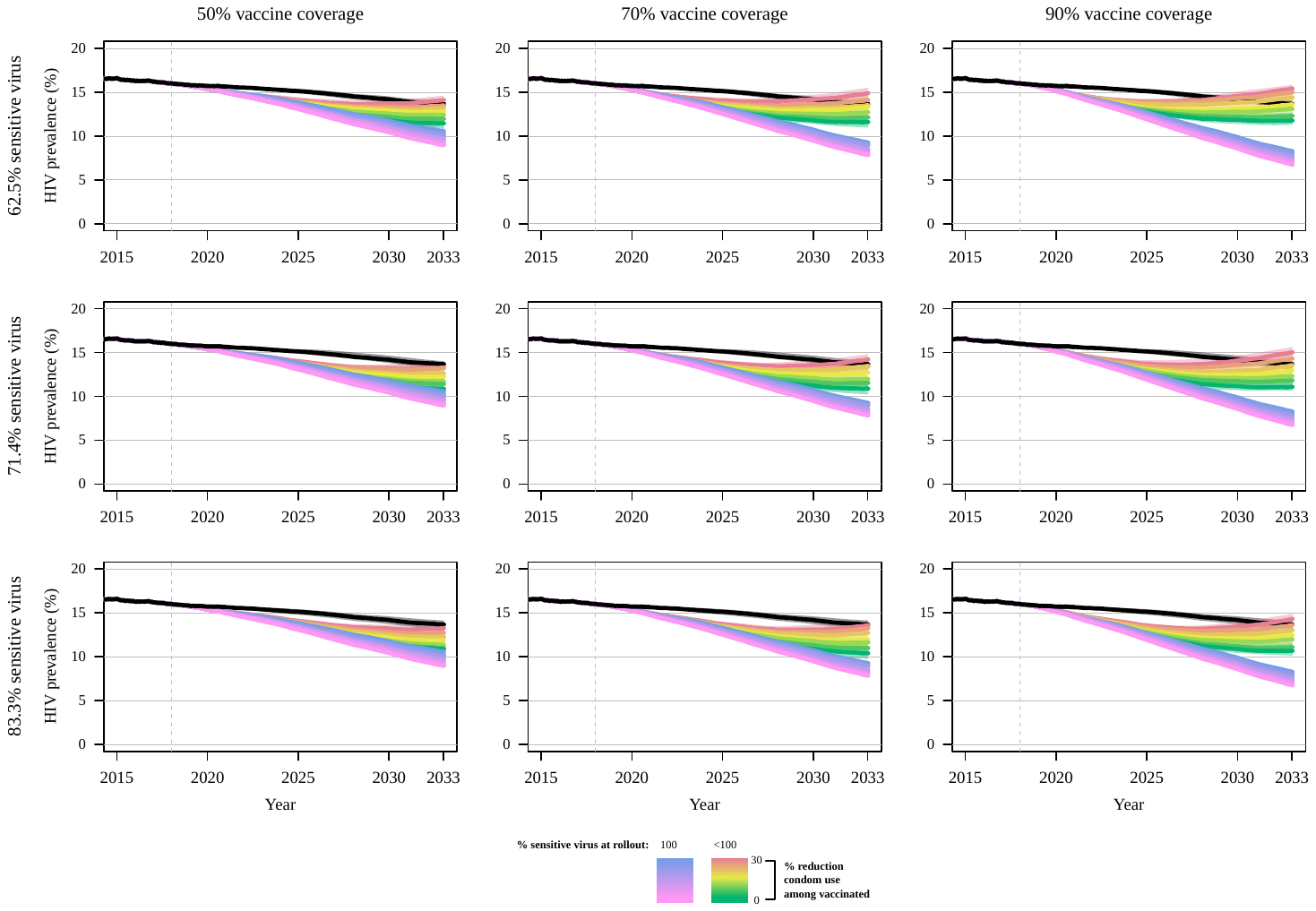

**Figure S5**. HIV-1 incidence of vaccine-sensitive and vaccine-resistant viral strains following introduction of a partially effective vaccine in the year 2018. Scenarios include variation in the proportion of vaccine-sensitive virus at vaccine rollout (62.5%, 71.4%, and 83.3% compared to 100%) and variation in the relative reduction in condom use among vaccinated individuals (from 0% to 30% reduction). The mean and 95% credible interval of 64 replicates of each scenario are shown.

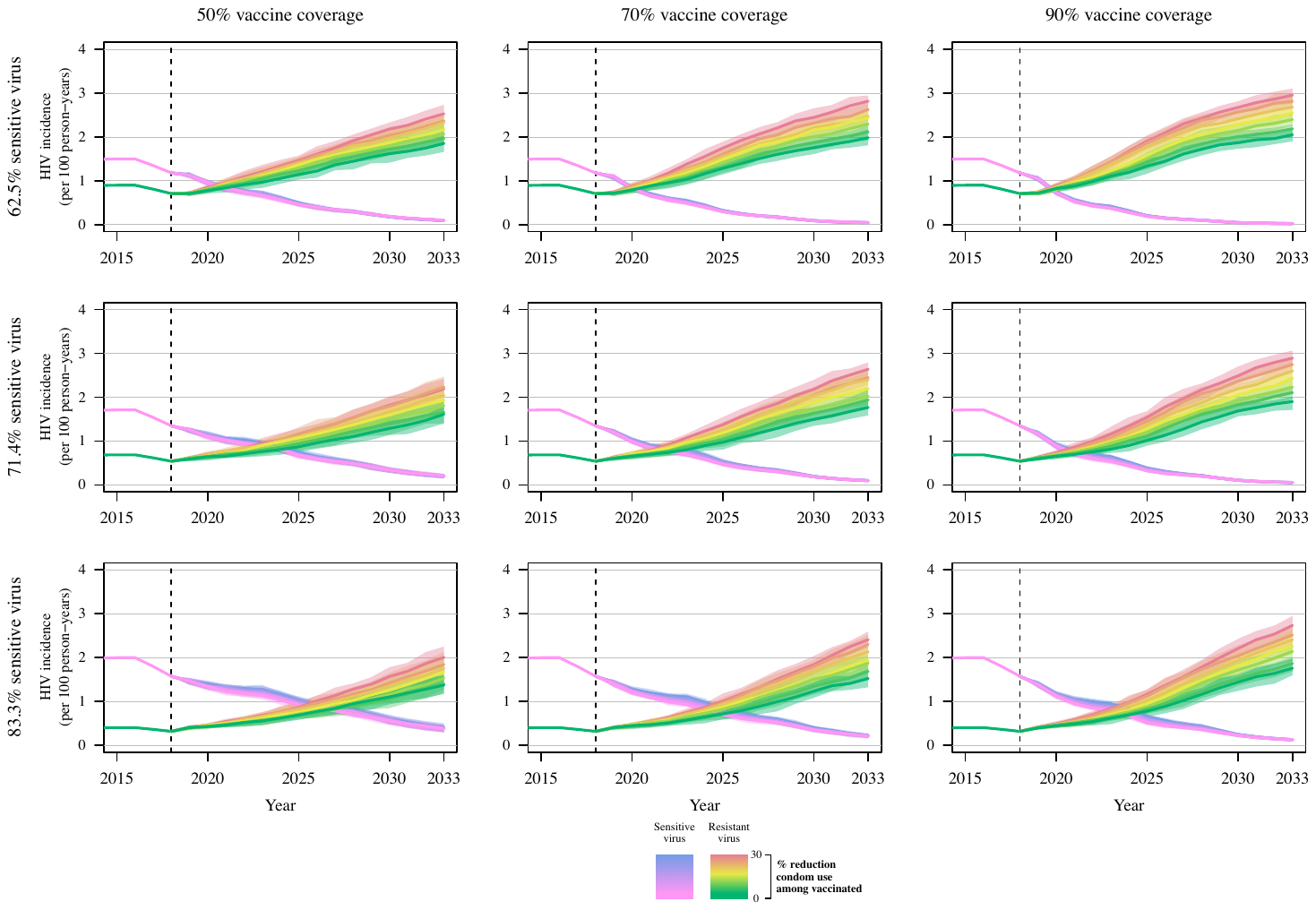

**Figure S6**. Percent of HIV-1 infections averted following 5, 10, and 15 years of continuous vaccine implementation. The mean and 95% credible interval of 64 replicates of each scenario are shown.

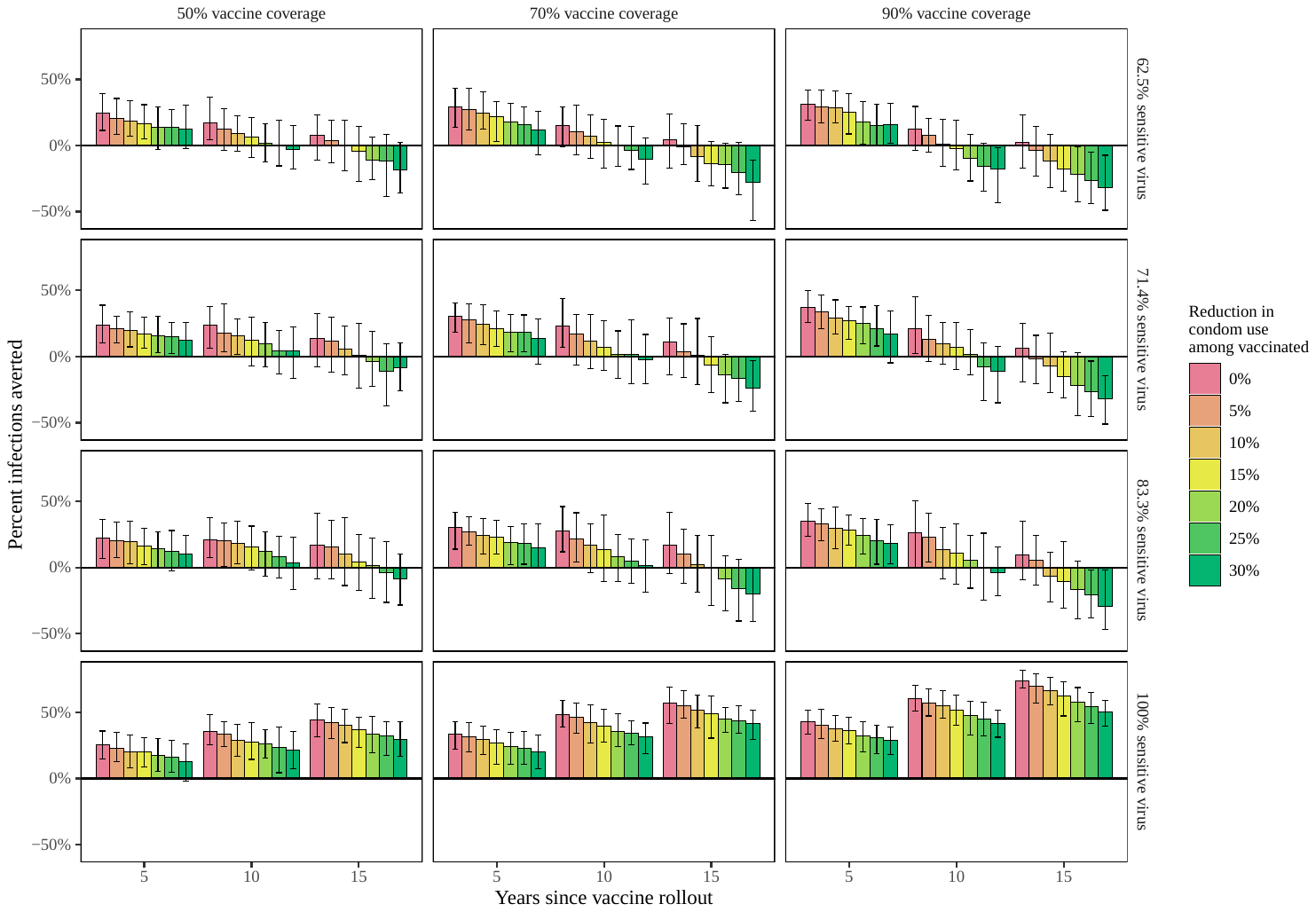

**Figure S7**. Incremental costs and infections averted following 15 years of continuous vaccine implementation. Health outcomes and costs are discounted at an annual rate of 3%. The mean and 90% credible interval of 64 replicates of each scenario are shown.

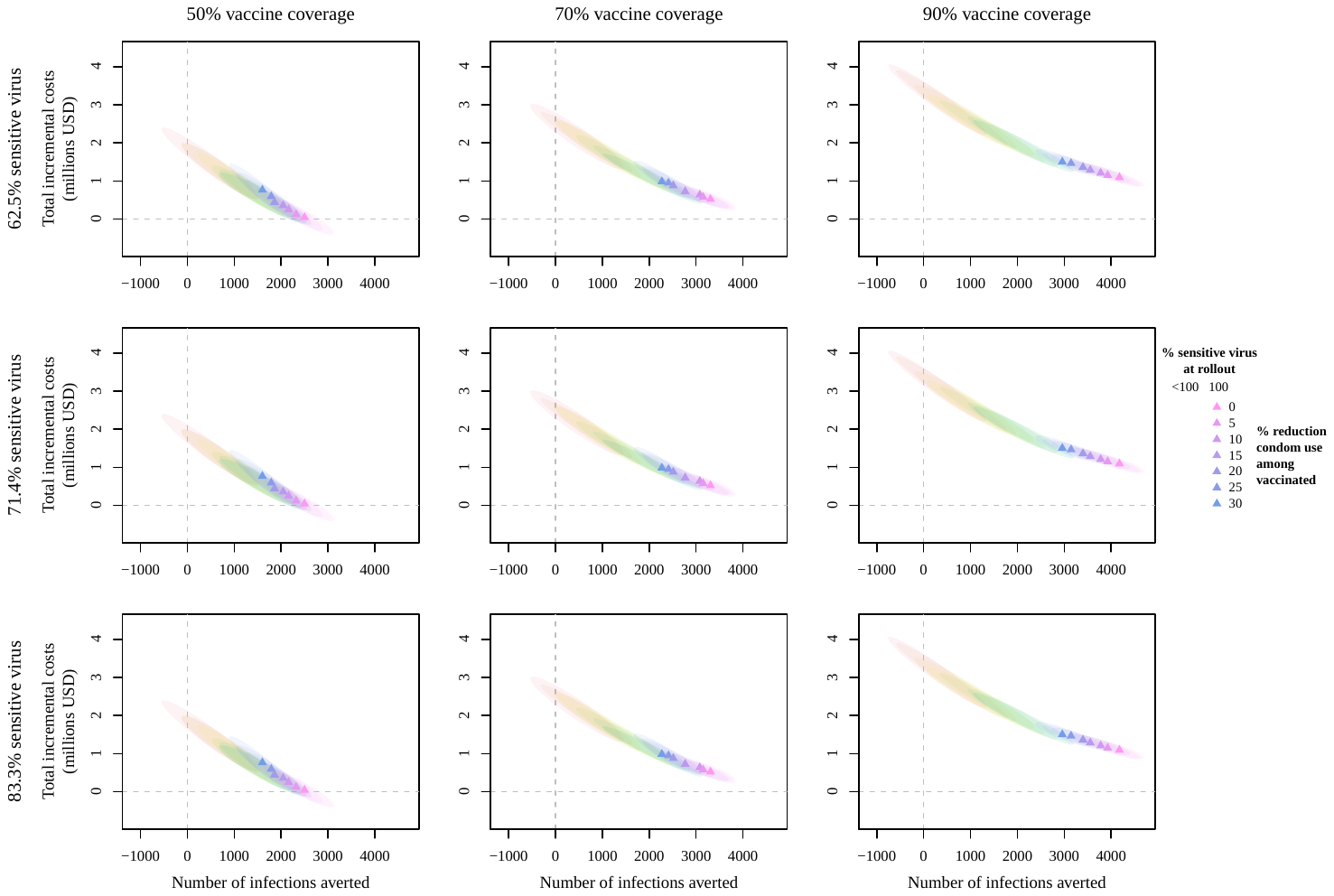

**Figure S8**. Incremental costs and disability-adjusted life-years (DALYs) averted following 15 years of continuous vaccine implementation. Health outcomes and costs are discounted at an annual rate of 3%. The mean and 90% credible intervals of 64 replicates of each scenario are shown.

**Figure S9**. Model-simulated proportion of condom-protected acts per time step and observed prevalence of condom use.

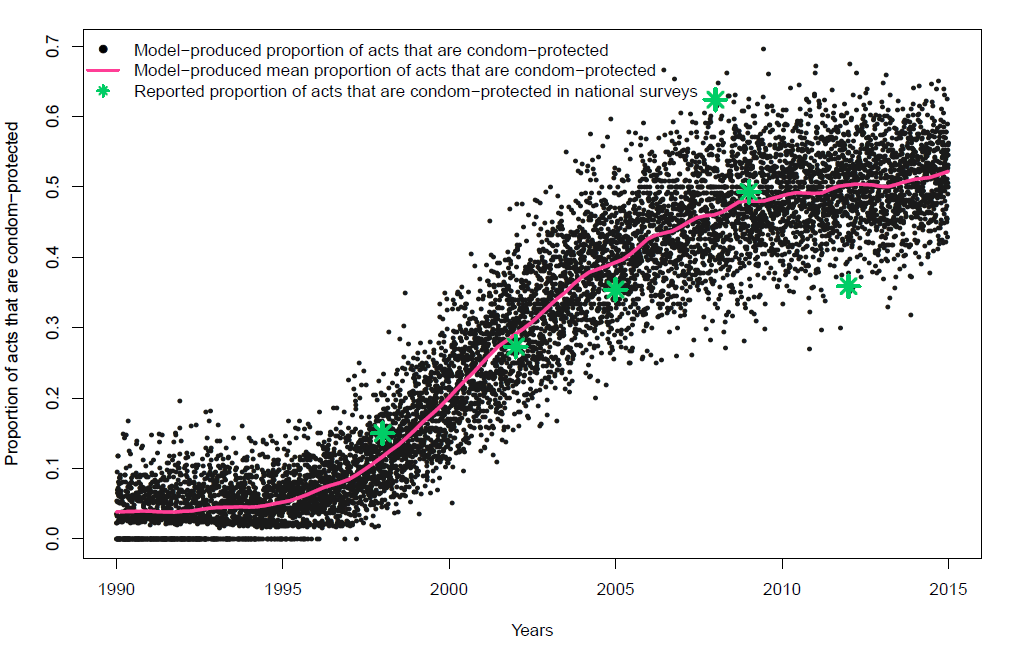

**Figure S10.** Intra-individual vaccine waning and boosting for vaccine efficacy at two years post-vaccination of 60%, 70%, and 80%

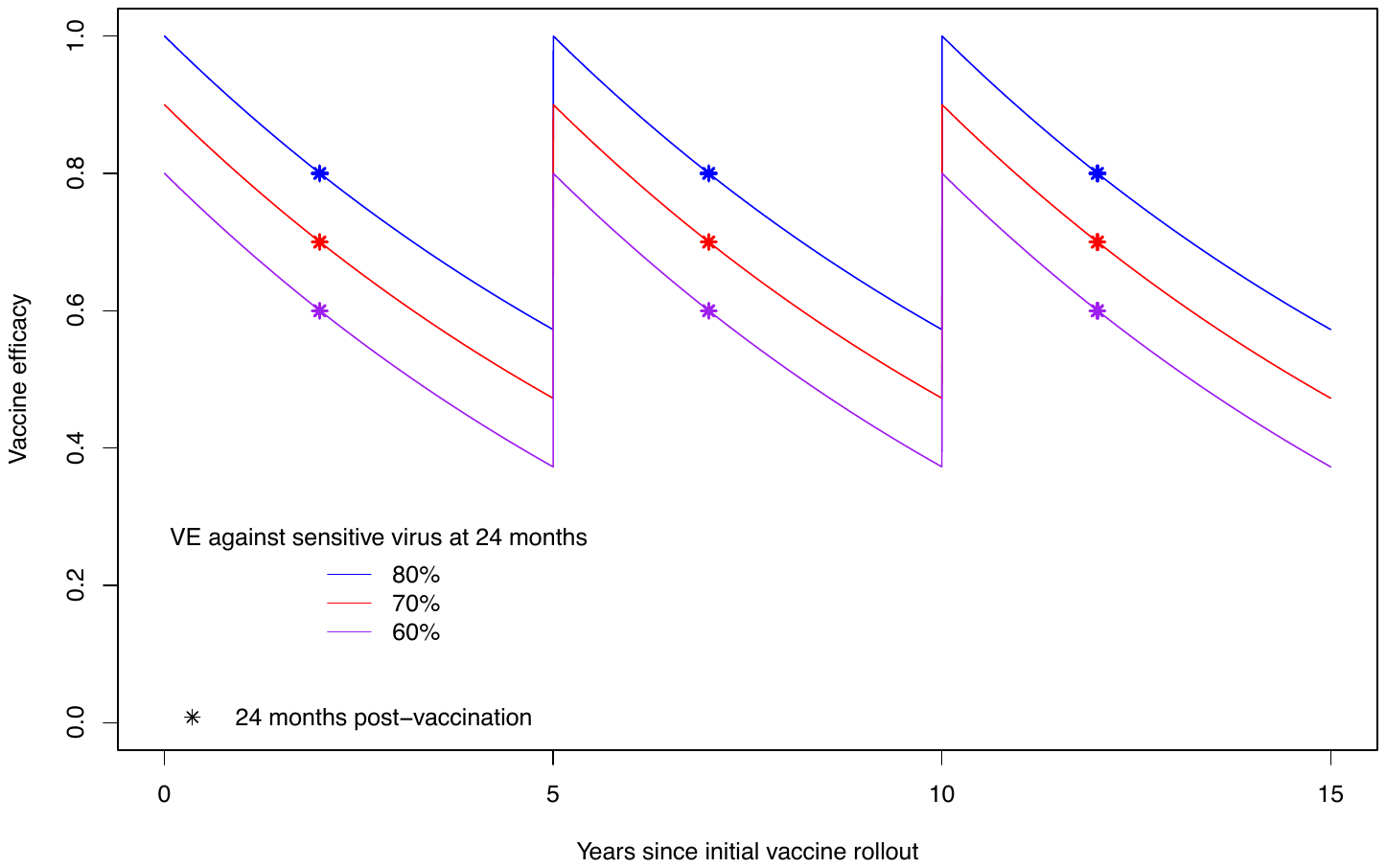
